# Influenza A Virus detection in Bulk Tank and Pen Level Milk from Dairies Affected by Highly Pathogenic Avian Influenza H5N1

**DOI:** 10.1101/2025.10.26.25338833

**Authors:** C. Stenkamp-Strahm, J. Lombard, B. Melody, P. Brinson, B. McCluskey

**Author notes:** Department of Veterinary Clinical and Life Sciences, College of Veterinary Medicine, Utah State University, Logan, UT 84322-1600. Lonestar Epidemiology Consulting LLC, 622 Linecamp Dr, Livermore, CO 80536. Corresponding author: Chloe Stenkamp-Strahm, 950 E 1400 N, Logan, UT 84341.

## Abstract

Highly pathogenic avian influenza virus H5N1 has been infecting dairy herds in the U.S. since its initial incursion into cows in early 2024. Although national strategies have aimed to detect affected herds, the best way to surveil herds for the H5N1 virus has not been formally studied and we also do not understand herd-level patterns of infection. To understand infection patterns of H5N1 in dairy herds over time, we conducted early surveillance of non-affected farms in California in the Fall of 2024 in an observational study. Daily bulk tank milk (BTM) samples were submitted from each herd and tested for influenza A (IAV) via rRt-PCR. In a subset of herds, IAV testing of multiple excretion types from cattle of different classes and pen-level daily milk was also completed soon after BTM detection. Daily detections of IAV occurred in BTM for a minimum of 33 days, with some herds continuing to have detection beyond a 75-day window. BTM Ct nadirs were seen between 1-3 weeks of detection. In herds that were tested, virus was detected in the milk from all pens of cattle within a very short time frame after BTM detection, or prior to the initiation of pen level sampling. A very low percentage (2.8%) of individual cow samples tested positive for IAV when collected soon after BTM detection, and although the virus was found in all excretion types, a majority of positive samples were from milk. This suggests that BTM may be the best early indicator of herd infection, and that movement of the virus to all lactating pens of cattle after herd incursion is relatively quick. These results also suggest that surveillance strategies with a long interval between BTM testing days may miss herds with short infection windows. Because most herds experienced test days where some submitted BTM samples had virus detected while others did not, and virus was detected in pen level milk samples when the BTM from the herd had become test negative, this work also highlights the necessity of studying the test sensitivity of IAV rRt-PCR detection in aggregate milk samples.

**Interpretive summary:** This study characterized daily bulk tank and pen-level milk detections of H5N1 virus on affected dairies. Viral loads in bulk tank samples peaked within three weeks and declined by two months, though some herds showed prolonged detection. Individual cow testing revealed that bulk tank milk testing is more sensitive for H5N1 early outbreak detection, and also identified the likely presence of non-clinically affected cows shedding virus. Pen-level sampling revealed rapid herd-level spread, emphasizing the need for frequent and comprehensive milk sampling to improve detection sensitivity and support effective outbreak management for H5N1 on dairy operations.

## INTRODUCTION

Highly pathogenic avian influenza virus of the H5N1 subtype has been infecting dairy cattle and spreading among dairy herds in the U.S since early 2024 (Nguyen et al., 2024). Prior to this outbreak, influenza A viruses (IAV) of the H5N1 subtype (hereafter H5N1) had infected wild birds, poultry, and other mammals and humans, but had not shown evidence of efficient mammal-to-mammal transmission (Gilbertson and Subbarao 2023; Tammiranta et al., 2023; Graziosi et al., 2024). Since the start of this outbreak, H5N1 has infected 1,080 dairy herds in 18 states (USDA, 2024a). Although the main modes of H5N1 transmission among cattle are unknown, the virus appears to move within and among herds proficiently.

Cattle that are exposed to H5N1 can show signs of clinical disease, although these vary in type and severity among individuals. During infection, cattle can shed H5N1 in different excretions, with high viral loads sometimes detected in the milk (Caserta et al., 2024). Bulk tank milk (BTM) has been historically used for surveillance of dairy pathogens, as these samples can represent the pooled milk from all lactating cows on an operation (Nobrega et al., 2023a; 2023b). Although the milk of individual sick cows is diverted from the saleable stream, H5N1 has been detected in BTM samples from herds both before and after the onset of clinical signs of H5N1 disease (NMPF, 2024; Spackman et al. 2024; Stenkamp-Strahm et al., 2025). Ideal surveillance techniques for H5N1 on dairy operations are still being refined, but finding H5N1 in the milk from infected cattle and the aggregate milk from affected herds has been leveraged in surveillance approaches during the current U.S. outbreak, and good detection sensitivity has been seen when using these methods (USDA, 2025). In the Spring of 2024, the United States Department of Agriculture (USDA) attempted to reduce interstate spread of the virus by implementing a federal order mandating that lactating cows have milk tested for IAV prior to interstate movement (USDA, 2024b). Testing for virus in BTM or silo milk samples has also been used for initial detection, quarantine, and/or continuous monitoring of H5N1 affected herds at the state (CDA, 2024; CDFA, 2024; UDAF, 2024) and now federal (USDA, 2024c) levels.

During these H5N1 surveillance activities, aggregate milk testing of affected herds has occurred on either a weekly, monthly, or more infrequent basis, with methodology of sample collection varying across states. The best methods for dairy H5N1 surveillance are unknown, in part because the patterns of herd-level H5N1 detections over time are not understood. To determine the patterns and longevity of BTM detections on H5N1 affected dairy operations, daily viral testing of samples taken throughout the outbreak on each farm is necessary. This would include sampling in the period prior to clinical disease detection in cows. Knowledge of these BTM detection patterns may reveal an ideal methodology for continued nation-wide surveillance of H5N1. It also serves to inform farms that have not yet experienced H5N1 infection, regarding the timeframe for disease management and quarantine, should they become infected in the future.

Transmission of the H5N1 virus among dairy farms and between individual dairy cows is still not understood. Routes of dairy cow H5N1 exposure may include oral, respiratory or teat end exposure via milking equipment. Once a farm is affected, individual pens of cattle may be exposed to virus at different rates, and/or by different routes. To the author’s knowledge aggregate milk testing for H5N1 done at the pen level (pen level milk; hereafter denoted as PLM) has not been previously performed in dairy herds affected during this outbreak. PLM testing done after H5N1 detections are made in BTM may inform patterns of intra-herd H5N1 movement and viral transmission within a dairy farm. Sampling of individual cattle after H5N1 detections are made in BTM, and prior to the onset of clinical signs in the herd, would also be advantageous to understand potential transmission routes of the virus into the herd.

To describe detections at the herd level and also inform intra-herd movement of dairy H5N1, this observational study included testing of daily BTM and PLM samples submitted from multiple California dairy herds (n = 19 and 5, respectively) both prior to and during H5N1 infection. A subset of these farms (n = 6) also allowed sampling of individual cows from multiple cattle classes soon after BTM detection. BTM, PLM, and individual cow samples were tested for influenza A virus (IAV). Results from BTM, PLM and individual cow sampling are used to describe patterns of dairy H5N1 detections seen at the herd level throughout each outbreak period. These results are then used to guide a discussion of both intra-herd viral transmission, and the best methods for future dairy H5N1 surveillance.

## MATERIALS AND METHODS

### Study Design

In the central valley region of California, a convenience sample of dairy operations (n = 19) that had not experienced H5N1 infection were recruited for enrollment in this observational study. This investigation was conducted by Lander Veterinary Clinic (LVC) personnel and included three overlapping studies focusing on IAV detection: a BTM detection study, a PLM detection study, and an individual cow detection study. All herds were enrolled between 9/10/2024 and 11/14/2024 and had from 600 to 3,050 lactating cows. Demographic characteristics of enrolled herds by study participation are described in Table 1. Farm size, parlor type, breeds and classes of cattle represented on each enrolled operation are described in Supplemental Table 1.

**Table 1:** Demographic characteristics of 19 California dairy herds enrolled in H5N1 detection studies, by study participation.

| Characteristic | Level | BTM Detection Study |  | PLM Detection Study |  | Individual Cow Detection Study |  |
| --- | --- | --- | --- | --- | --- | --- | --- |
|  |  | Number | Percent | Number | Percent | Number | Percent |
|  | Total | 19 | 100.0 | 5 | 100.0 | 6 | 100.0 |
| Herd Size | <1,000 | 5 | 26.3 | 0 | 0.0 | 1 | 16.7 |
|  | 1,000 - ≤2,000 | 7 | 36.8 | 3 | 60.0 | 3 | 50.0 |
|  | >2,000 | 7 | 36.8 | 2 | 40.0 | 2 | 33.3 |
| Times milked per day | ≤ 2 | 4 | 21.1 | 1 | 20.0 | 1 | 16.7 |
|  | 2 – 3<sup>1</sup> | 4 | 21.1 | 2 | 40.0 | 2 | 33.3 |
|  | ≥ 3 | 11 | 57.8 | 2 | 40.0 | 3 | 50.0 |
| Breed | Holstein ≥95% | 13 | 68.4 | 3 | 60.0 | 5 | 83.3 |
|  | Jersey ≥95% | 2 | 10.5 | 2 | 40.0 | 1 | 16.7 |
|  | Mixed | 4 | 21.1 | 0 | 0.0 | 0 | 0.0 |
| Parlor Type | Herringbone | 8 | 42.1 | 2 | 40.0 | 1 | 16.7 |
|  | Parallel | 4 | 21.1 | 1 | 20.0 | 1 | 16.7 |
|  | Rotary | 5 | 26.3 | 2 | 40.0 | 3 | 50.0 |
|  | Robotic | 1 | 5.3 | 0 | 0.0 | 1 | 16.7 |
|  | Flat Barn | 1 | 5.3 | 0 | 0.0 | 0.0 | 0.0 |

<sup>1</sup>Farm will milk cattle either 2 or 3 times per day, depending on production

For the BTM detection study, each operation submitted daily BTM samples starting at enrollment and continuing until 2/11/25. A single farm stopped daily submissions after 1/10/25, given their earlier outbreak timeframe, and lack of detection in submitted samples for the prior three weeks. Farms were instructed to submit one aggregate milk sample from each tank of shipped milk. Farms submitted between one and eight unique BTM samples per day depending on operation size and daily milk production. For the PLM detection study, a subset of farms (n = 5) submitted pooled saleable milk from each pen of lactating cows daily, beginning six to 18 days after initial BTM detection of IAV. For the individual cow detection study, six operations allowed sampling of cattle within nine days of BTM detection. Samples were collected from lactating cows that had clinical signs of either H5N1 or other diseases (sick cows), non-clinically affected lactating cows (healthy cows), dry cows (non-lactating cows), fresh cows (those less than 21 days post-calving) pre-weaned and weaned heifers. Samples collected included serum, nasal swabs, urine, and a composite milk sample from all functional quarters if lactating. Milk from individual quarters was collected from a subset of 20 sick cows on a single operation.

Individual cow records were collected from on-farm record management systems. Dates that each farm first experienced clinical signs were determined using herd level records, and discussions with veterinarians and herd management personnel.

### Sample Collection and Processing

On each farm, milk haulers and dairy staff were trained by LVC personnel to collect BTM and PLM samples, respectively, and these were kept at 0°C until courier pick-up. Farms were instructed to collect one sample from each filled bulk tank. For PLM collection, standard operating procedures were followed using a QualiTru Sampling Systems peristaltic pump (Part No. 500200) with a sanitary fitting, a sterile seven channel septum, and sterile collection bags. Between each milking pen, a new sterile septa channel and fluid line were fitted and residual milk was flushed from the receiver jar(s) several times to allow it to clear prior to subsequent collection. Farms enrolled in this portion of the project submitted once daily PLM samples from all lactating pens of cattle on the operation (n = 6 – 13) that contributed saleable milk to the farm bulk tank(s). Couriers collected BTM and PLM samples from each farm daily. Samples were driven to LVC on ice and refrigerated on arrival.

BTM and PLM samples were characterized for IAV via rRT-PCR assays at LVC within 24 hours of collection. The following protocol was used for RNA isolation and subsequent PCR: RNA extraction was completed using 50ul of sample diluted in 150ul phosphate buffered saline with a MagMax CORE Nucleic Acid Purification Kit (Applied Biosystems, Cat. A32700) paired with the Swine Influenza Virus RNA Test Kit (Applied Biosystems, Cat. 4415200) on a KingFisher Flex 96 system (ThermoFisher). A QuantStudio 6 or QuantStudio 5 Pro Real-Time PCR System (ThermoFisher, Cat. A43168, A28569) was used according to manufacturer’s instructions to perform rRT-PCR for IAV. Reactions were performed using 8ul of extracted RNA from each sample. A cycle threshold (Ct) cut-off value of 40 was used. Samples were stored at 4°C after testing.

For the six operations enrolled to sample individual cattle, attempts were made on each farm to collect samples from >= 20 sick lactating cows, 10-15 healthy lactating cows, nonlactating cows, cows that had recently calved (fresh cows), and preweaned and/or weaned calves. Collection of samples was dependent on which cattle classes were present on each operation. Cattle classified as sick were selected by farm personnel or were cattle present in a hospital pen due to various clinical diseases (e.g., lameness, lethargy, gastrointestinal disease, pneumonia, and suspected H5N1). Milk from individual cows was collected in the parlor after teats had been prepped and prior to the placement of milking units, or when cows were restrained in housing areas, and placed in 50ml conical vials. A single operation had milk samples collected from individual quarters on a subset of cattle (n = 20), otherwise milk was collected as a composite of milk from all four quarters. When cows were restrained in housing areas, urine was collected in 50ml conical tubes, blood was collected from the coccygeal vein into serum separator tubes, and nasal swab samples were collected from both nostrils using sterile swabs. Swabs were placed in 3-5ml transport media (brain heart infusion (BHI) or molecular transport media (MTM)) provided by USDA’s National Veterinary Services Laboratories (NVSL, Ames, IA). All samples from individual cattle were kept on ice after collection.

BTM, PLM and all individual cow samples were shipped to a National Animal Health Laboratory Network (NAHLN) lab; Iowa State University (ISU) Veterinary Diagnostic Laboratory, in Ames, IA. Sample RNA was isolated and rRT-PCR assays for IAV were performed per NAHLN standard operating procedures and protocols, using a cycle threshold (Ct) cut-off value of 40. Ct values <40 were considered positive. Annually, each NAHLN laboratory is required to successfully complete NVSL-administered proficiency tests and quality control procedures for identification of IAV. Serum samples from cattle were additionally tested using an IDEXX AI MultiS-Screen ELISA (IDEXX, Cat. 99-12119; Westbrook, ME) per NAHLN standard operating procedures and protocols. Samples were determined as ELISA positive if the signal to noise (S/N) ratio was ≤ 0.4. Samples were considered suspect positive at S/N ratios > 0.4 and < 0.5, and negative if ratios were ≥ 0.5.

After initial detection of IAV in the BTM from each operation, each herd was confirmed to be infected with IAV H5N1 clade 2.3.4.4b genotype B3.13 via testing at NVSL.

### Analysis

BTM samples on some farms were not collected every day. Dates with missing IAV results from enrolled farms (Supplementary Table 2) represented weekend or holiday dates where sample collection was not possible, or dates where shipments of samples were either misplaced or lost in transit. A majority of rRT-PCR results used in subsequent analyses were from testing at the ISU NAHLN laboratory. Supplementary Table 2 describes dates where sample Ct values were supplied from rRT-PCR assays done at LVC due to shipments of samples having been misplaced, or changes in study funding source. To ensure comparable test results, a correlation of Ct values from LVC and ISU was completed by selecting dates from 5 farms with low (< 25), medium (25 – 35) and high (> 35) Ct values measured at each testing lab, for comparison. Pearson correlation and Wilcoxon signed rank for paired sample tests were performed using these values, and a correlation regression plot was created (Supplemental Figure 1).

For each farm, the number of days from BTM detection to the onset of clinical signs in cows, the number of days from BTM detection or onset of clinical signs to the lowest achieved BTM Ct value, and the number of days from BTM detection to the first day all samples submitted were non-detected, were calculated. The lowest BTM and PLM Ct value measured for each farm over the course of the study was determined, and the lowest daily Ct values measured for BTM and PLM was graphically depicted over time. The lowest Ct value from each pen was described by the time from initial BTM detection and the onset of clinical signs in cows. The number of days that each farm submitted milk samples with both a detected and non-detected delineation was also calculated. For individual cows, the number of samples with IAV detected was described by cattle class and sample type on each operation. Statistical tests were performed using R version 2024.12.1+563 or later.

## RESULTS

Of operations enrolled, a majority had more than 1,000 lactating cows (73.6% for the BTM study, 100% for the PLM study, and 83.3% for the individual cow study) (Table 1). About half of operations milked their lactating cows three times per day (57.8% for the BTM study, 40% for the PLM study, and 50% for the individual cow study). For the BTM study, 68.4% of the operations were comprised of primarily Holstein cows, while 10.5% had primarily Jersey cows and the other 21.1% of farms were composed of multiple or mixed breeds. For the PLM and individual cow detection studies, most operations were comprised of Holsteins (60.0% and 83.3% for PLM and individual cow detection, respectively), while the rest were comprised of Jersey cows (40.0% and 16.7%, for PLM and individual cow, respectively). Operations enrolled across studies had several different parlor types, including herringbone, parallel, rotary, robotic and flat barn (Table 1). The single highest percentage of herds enrolled in the BTM study had herringbone parlors (42.1% compared to 21.1%, 26.3%, 5.3%, 5.3% for parallel, robotic, rotary and flat barn parlors, respectively) while an equal number of PLM herds had herringbone and rotary parlors (40% for herringbone and rotary, compared to 20% parallel, respectively). Half of the farms (50%) in the individual cow detection study had rotary parlors compared to 16.7% each for herringbone, parallel, and robotic parlor types.

During the study period, a total of 7,671 aggregate milk samples were submitted from enrolled operations; 4,749 were BTM and 2,922 were PLM. The total number of BTM and PLM samples submitted by each farm can be found in Supplemental Table 1. A total of 16 farms submitted BTM samples consistently throughout the study period with the remaining 3 farms being omitted from subsequent analyses due to lack of, or inconsistencies with, daily sample submission. All other farms submitted samples daily, with some exceptions (see missing submissions, Supplemental Table 2). Although samples were characterized for IAV at both NAHLN and LVC laboratories (Supplemental Table 2; percentage LVC results), a correlation using sample results from five study farms showed a high correlation (Pearson = 0.954; Supplemental Figure 1) and lack of significant difference (Shapiro-wilk p<0.001; Wilcoxon signed-rank = 0.796) between values. Of note, most study farms experienced days where some BTM samples submitted had IAV detected, while other BTM samples submitted had IAV non-detected (range in number of days with detection and non-detection = 0 – 19; Supplemental Table 2).

### Bulk Tank Milk Detection

Each enrolled farm had initial BTM IAV detection between 10/24/24 and 12/11/24. A timeline showing initial BTM detection, first clinical signs of H5N1 disease in cows, the first date that all samples submitted from each farm after detection were negative, and/or the end date of sampling can be seen in Figure 1. The number of days from initial IAV detection to the first negative BTM sample on each farm ranged from 33 to 77 days, with three farms having continuous daily detections until the end of the study period (Table 2, Figure 1). Of note, three farms were missing BTM sample submissions in the days leading up to first detection; these were not included in the Figure 1 timeline or the BTM detection length calculations in Table 2.

**Figure 1:**
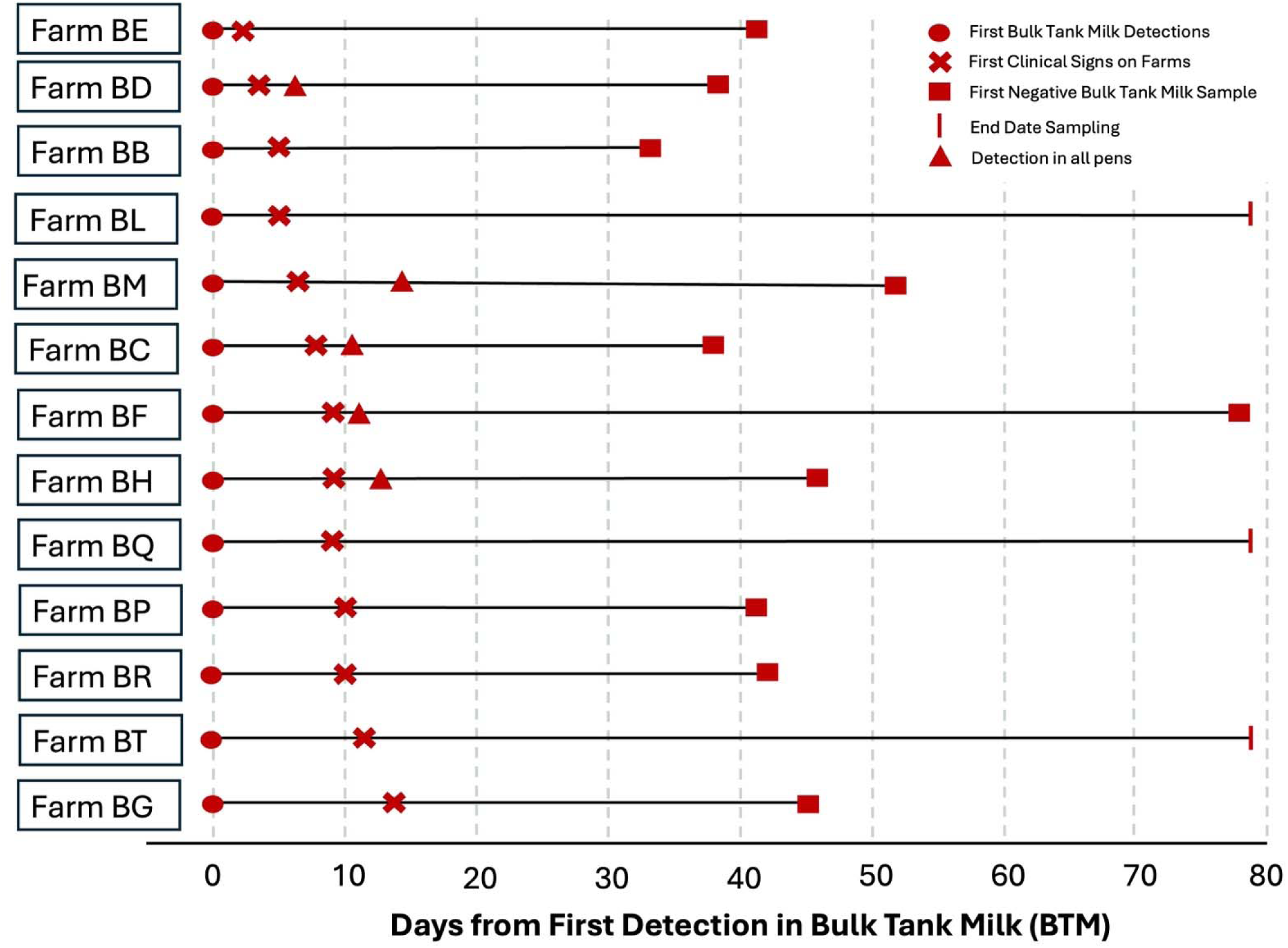
Timeline showing the number of days from BTM influenza A virus (IAV) detection to clinical signs in 13 California dairy herds, IAV detection in PLM from all pens (for farms enrolled in the PLM study), and date of either the first day with all negative tanks or end date of sampling. Study herds shown had consistent daily submissions of BTM throughout the study period.

**Table 2:**
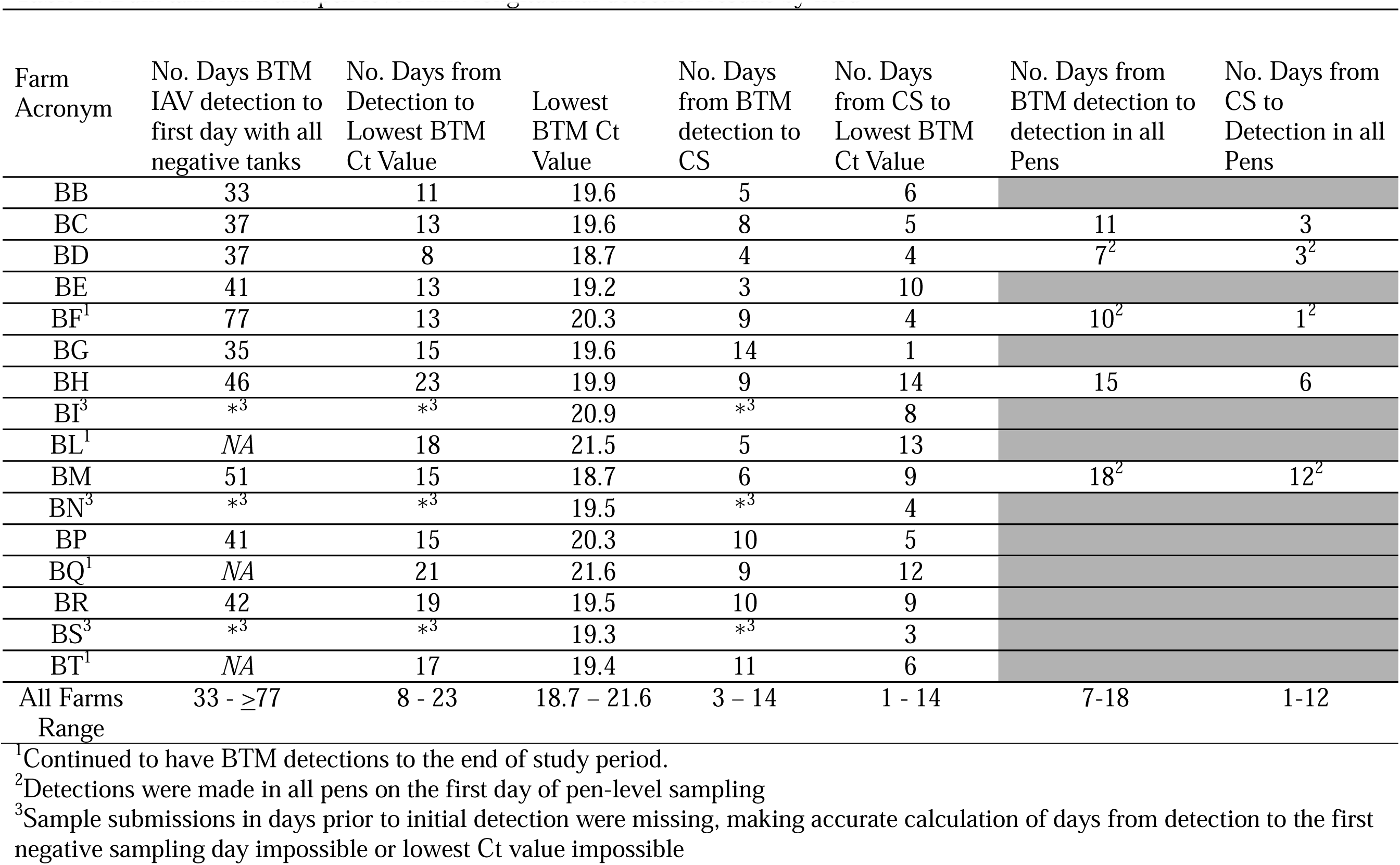
Bulk tank milk and pen level milk longitudinal detection results by herd.

| Farm Acronym | No. Days BTM IAV detection to first day with all negative tanks | No. Days from Detection to Lowest BTM Ct Value | Lowest BTM Ct Value | No. Days from BTM detection to CS | No. Days from CS to Lowest BTM Ct Value | No. Days from BTM detection to detection in all Pens | No. Days from CS to Detection in all Pens |
| --- | --- | --- | --- | --- | --- | --- | --- |
| BB | 33 | 11 | 19.6 | 5 | 6 |  |  |
| BC | 37 | 13 | 19.6 | 8 | 5 | 11 | 3 |
| BD | 37 | 8 | 18.7 | 4 | 4 | 7<sup>2</sup> | 3<sup>2</sup> |
| BE | 41 | 13 | 19.2 | 3 | 10 |  |  |
| BF<sup>1</sup> | 77 | 13 | 20.3 | 9 | 4 | 10<sup>2</sup> | 1<sup>2</sup> |
| BG | 35 | 15 | 19.6 | 14 | 1 |  |  |
| BH | 46 | 23 | 19.9 | 9 | 14 | 15 | 6 |
| BI<sup>3</sup> | *<sup>3</sup> | *<sup>3</sup> | 20.9 | *<sup>3</sup> | 8 |  |  |
| BL<sup>1</sup> | NA | 18 | 21.5 | 5 | 13 |  |  |
| BM | 51 | 15 | 18.7 | 6 | 9 | 18<sup>2</sup> | 12<sup>2</sup> |
| BN<sup>3</sup> | *<sup>3</sup> | *<sup>3</sup> | 19.5 | *<sup>3</sup> | 4 |  |  |
| BP | 41 | 15 | 20.3 | 10 | 5 |  |  |
| BQ<sup>1</sup> | NA | 21 | 21.6 | 9 | 12 |  |  |
| BR | 42 | 19 | 19.5 | 10 | 9 |  |  |
| BS<sup>3</sup> | *<sup>3</sup> | *<sup>3</sup> | 19.3 | *<sup>3</sup> | 3 |  |  |
| BT<sup>1</sup> | NA | 17 | 19.4 | 11 | 6 |  |  |
| All Farms Range | 33 - ≥77 | 8 - 23 | 18.7 – 21.6 | 3 – 14 | 1 - 14 | 7-18 | 1-12 |

<sup>1</sup>Continued to have BTM detections to the end of study period.
<sup>2</sup>Detections were made in all pens on the first day of pen-level sampling
<sup>3</sup>Sample submissions in days prior to initial detection were missing, making accurate calculation of days from detection to the first negative sampling day impossible or lowest Ct value impossible

The lowest BTM Ct value achieved during the outbreak period on each enrolled farm was between 18.7 and 21.6 and occurred from eight to 23 days after initial BTM detection (Table 2). Each farm experienced clinical signs of H5N1 disease in cows between three and 14 days of initial BTM detection, and clinical signs occurred one to 14 days prior to each farm’s lowest achieved BTM Ct (Table 2). Curves of the lowest daily BTM Ct values over time for six representative farms can be seen in Figure 2, with curves from the additional ten study farms found in Supplemental Figure 2.

**Figure 2:**
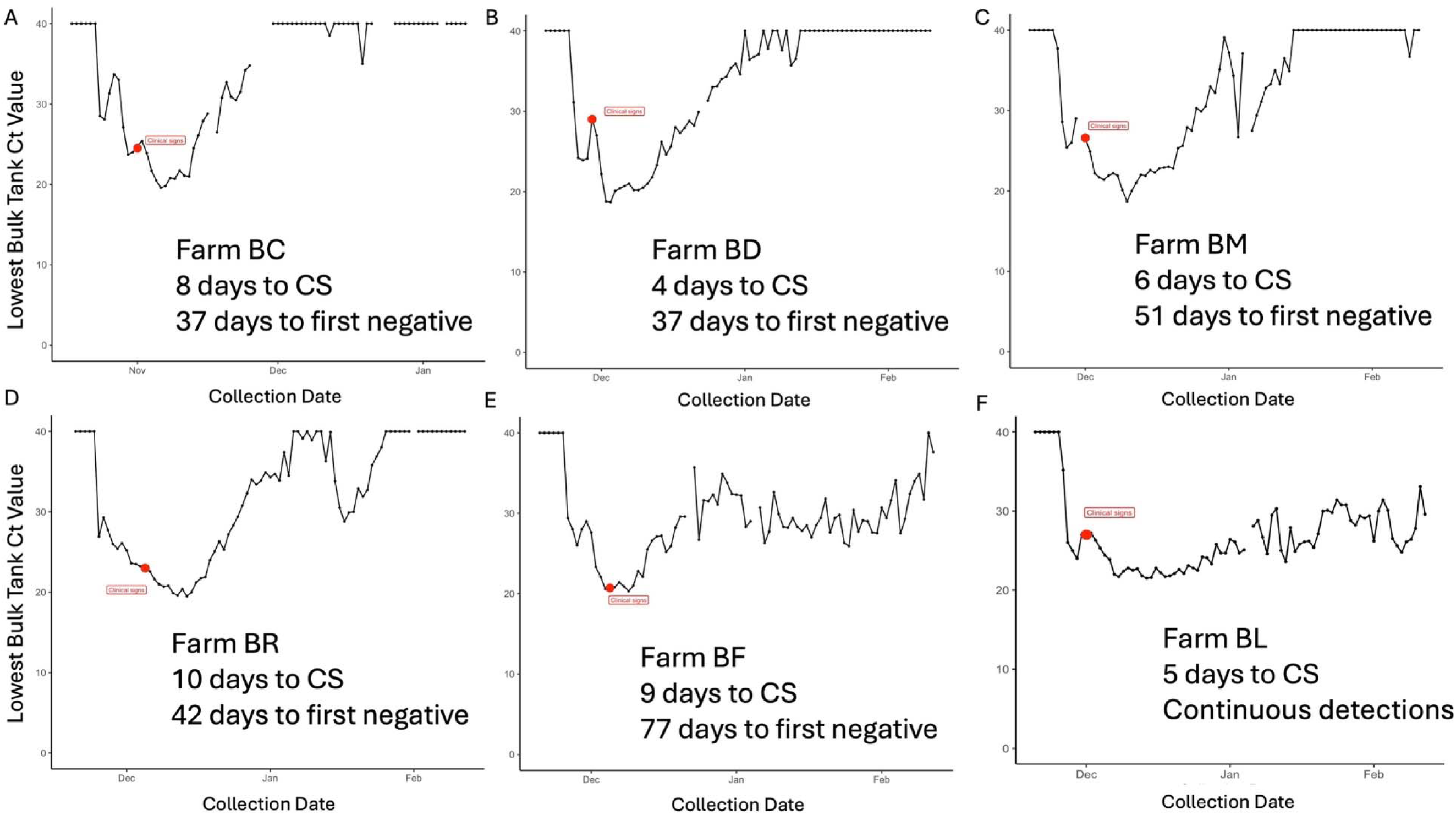
Panels A-F: Lowest daily influenza A (IAV) Ct values from bulk tank milk (BTM) samples submitted from six (6) California dairy farms over time. These farms had a different number of days from BTM IAV detection to when cows experienced clinical signs (CS), and different periods of time (37 – 77 days) between initial IAV detection to when all tank samples submitted on a day were negative. Two of the farms represented (Panel E, Panel F) continued to have BTM IAV detections to the end of the study period.

### Pen Level Milk Detection

PLM samples were collected on five enrolled farms starting six to 18 days after each farm had initial IAV detection in their BTM. IAV was detected in the milk from all pens on these operations seven to 18 days of initial BTM detection, although three of five farms had detection in all pens on their first pen-level sample collection date (at 7-, 10-, and 18-days post BTM detection, respectively; Table 2, Figure 1). The number of days from clinical signs to IAV detection in all PLM samples ranged from one to 12 days for these farms (Table 2, Figure 1), although these estimates could be erroneously long given the delay in starting PLM sample collection. Curves showing daily PLM Ct values with daily BTM Ct values from each of the five represented farms are seen in Figure 3. PLM samples collected near the time of the BTM Ct nadir on each farm tended to have Ct values similar to those of the main bulk tank (Figure 3). Of note, there were days when PLM IAV detections were made when BTM from the same herd were test-negative for IAV (Figure 3B, 3D). A single farm that submitted PLM samples continued to have BTM detections until the end of the study period (Figure 3C). In the final days of sampling, only two pens appeared to be contributing to continued detections on this farm. Cattle in these pens included cows and heifers that had recently calved (i.e., fresh cows), and high producing cows in their first to fourth lactations.

**Figure 3:**
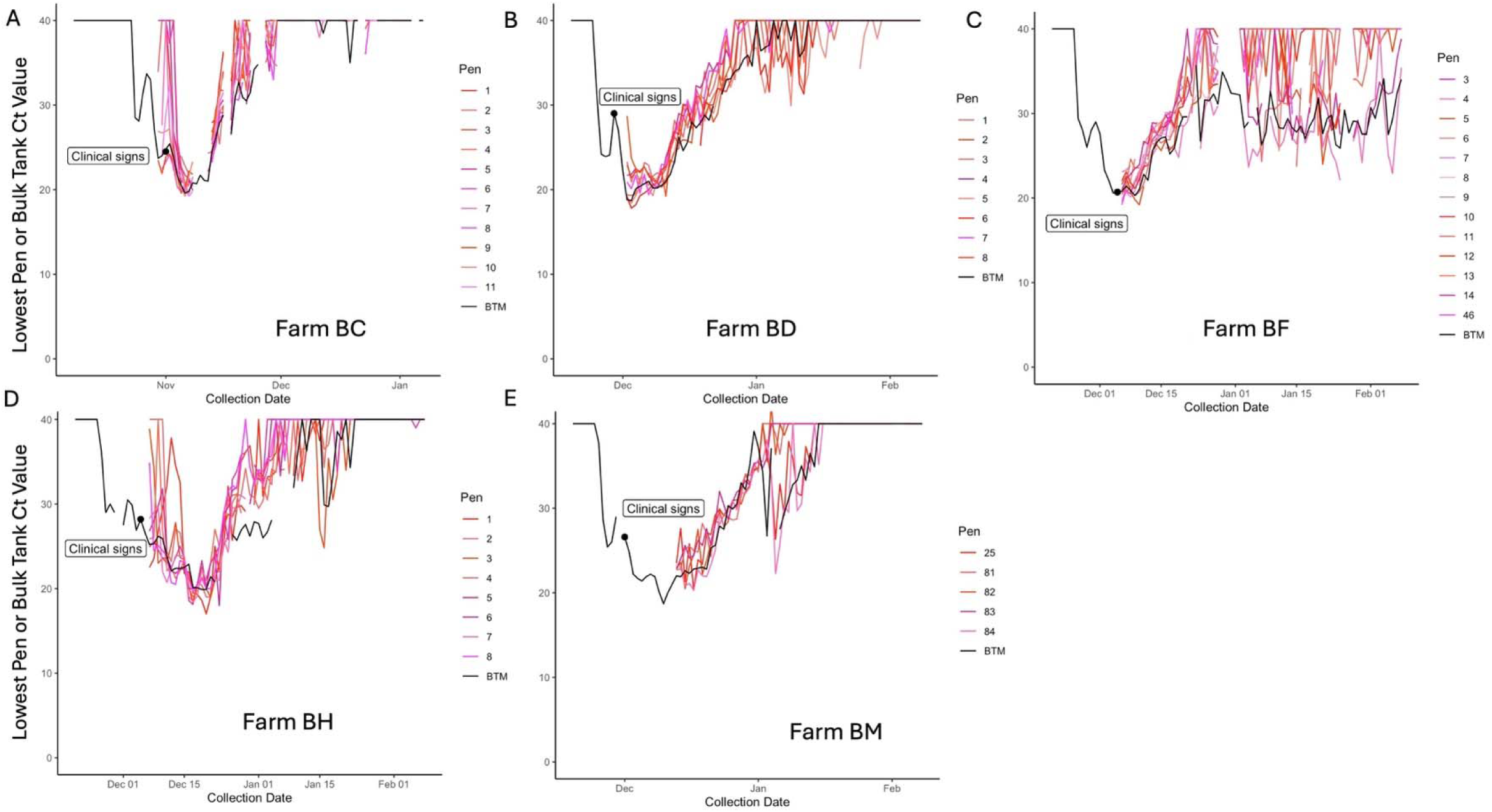
Panels A-E: Lowest daily influenza A (IAV) Ct values from bulk tank milk or pen-level milk samples submitted from five California dairy farms enrolled in the pen-level detection study, over time. Some farms (Panels B, C, E) had detection in all pens on the first day pen-level samples were submitted. One farm (Panel C) had detections in BTM and some pen-level samples to the end of the study period. One farm (Panel B) had several days where detections were made in pen-level milk samples even though BTM had become test-negative.

### Individual Cow Sampling

A total of 1,309 samples of milk, nasal swabs, serum and urine were collected from individual cows on six enrolled farms (Farms BC, BD, BI, BL, BM, and BN) four to nine days after BTM detection (Table 3). Of these samples, 419 came from sick cows, 198 came from healthy cows, 223 from nonlactating cows, 245 from fresh cows, 152 from preweaned calves and 72 from weaned calves (Table 3). Of all individual cow samples collected, just 2.8% (37) had IAV detected, representing 33 cows. The highest percentage of positive samples were taken from sicks cows (8%) followed by healthy cows (1%) and fresh cows (0.4%) (Table 3). No samples from non-lactating cows, preweaned or weaned calves had IAV detected. All cattle had detection in a single excretion, except for two sick cows on Farm BN that had detections in both milk and urine, and one sick cow on Farm BD that had detections in milk, urine, and serum. At the farm level, Farm BN had the highest percentage of samples with detection (11%) followed by Farm BD and Farm BM (4% for each Table 3). Farm BC, Farm BI and Farm BL each had less than 1% of samples with detection. For specific sample types, milk samples had the highest percent with detection (12.7%), followed by urine (2%) (Table 4). Nasal swab and serum samples had one detection each (0.3% for each; Table 4). Of note, quarter samples collected from 20 sick cows on Farm BC were all negative; the single PCR-positive milk sample from this operation was a composite sample.

**Table 3:** Summary of individual cow sample Influenza A virus PCR-positive test results for 6 California dairy herds/farms, by cattle class and farm of collection.

| Sample type | Number of PCR Positive Samples by Cattle Class |  |  |  |  |  | Total |  |
| --- | --- | --- | --- | --- | --- | --- | --- | --- |
|  | Sick Cows | Healthy Cows | Non-lactating Cows | Fresh Cows | Preweaned Heifers | Weaned Heifers | Number | Percent |
| **Farm BC (5-6<sup>1</sup>)** |  |  |  |  |  |  |  |  |
| Milk<sup>2</sup> | 1/25 | 0/11 |  | 0/15 |  |  | 1/53 | 1.9% |
| Nasal Swab | 0/24 | 0/13 | 0/15 | 0/15 | 0/11 | 0/9 | 0/89 | 0% |
| Serum | 0/23 | 0/11 | 0/15 | 0/15 | 0/6 | 0/9 | 0/79 | 0% |
| Urine | 1/24 | 0/11 | 0/14 | 0/12 |  | 0/9 | 1/70 | 1.4% |
| Total | 2/96 | 0/46 | 0/44 | 0/57 | 0/17 | 0/27 | 2/287 | 0.7% |
| **Farm BD (5<sup>1</sup>)** |  |  |  |  |  |  |  |  |
| Milk | 9/24 | 0/10 |  | 0/13 |  |  | 9/47 | 19% |
| Nasal Swab | 0/24 | 0/10 | 0/15 | 0/14 |  | 0/15 | 0/78 | 0% |
| Serum | 1/24 | 0/10 | 0/15 | 0/13 |  | 0/15 | 1/77 | 1.3% |
| Urine | 1/20 | 0/10 | 0/15 | 0/13 |  | 0/15 | 1/73 | 1.4% |
| Total | 11/92 | 0/40 | 0/45 | 0/53 |  | 0/45 | 11/275 | 4.0% |
| **Farm BI (4-7<sup>1</sup>)** |  |  |  |  |  |  |  |  |
| Milk |  |  |  | 0/9 |  |  | 0/9 | 0% |
| Nasal Swab |  |  | 0/15 | 0/9 | 0/15 |  | 0/39 | 0% |
| Serum |  |  | 0/14 | 0/9 | 0/15 |  | 0/38 | 0% |
| Urine |  |  | 0/15 | 1/9 | 0/15 |  | 1/39 | 2.6% |
| Total |  |  | 0/44 | 1/36 | 0/45 |  | 1/125 | 0.8% |
| **Farm BL (7<sup>1</sup>)** |  |  |  |  |  |  |  |  |
| Milk | 1/20 | 0/10 |  | 0/15 |  |  | 1/45 | 2.2% |
| Nasal Swab | 0/20 | 0/10 | 0/15 | 0/15 | 0/15 |  | 0/75 | 0% |
| Serum | 0/20 | 0/10 | 0/15 | 0/15 | 0/15 |  | 0/75 | 0% |
| Urine | 0/13 | 0/10 | 0/15 | 0/15 | 0/15 |  | 0/68 | 0% |
| Total | 1/73 | 0/40 | 0/45 | 0/60 | 0/45 |  | 1/263 | 0.4% |
| **Farm BM (9<sup>1</sup>)** |  |  |  |  |  |  |  |  |
| Milk | 9/21 | 0/9 |  | 0/10 |  |  | 9/40 | 23% |
| Nasal Swab | 1/21 | 0/9 | 0/15 | 0/10 | 0/15 |  | 1/70 | 1.4% |
| Serum | 0/21 | 0/9 | 0/15 | 0/10 | 0/15 |  | 0/70 | 0% |
| Urine | 0/21 | 0/9 | 0/15 | 0/9 | 0/15 |  | 0/69 | 0% |
| Total | 10/84 | 0/36 | 0/45 | 0/39 | 0/45 |  | 10/249 | 4.0% |
| **Farm BN (7<sup>1</sup>)** |  |  |  |  |  |  |  |  |
| Milk | 6/19 | 2/9 |  |  |  |  | 8/28 | 29% |
| Nasal Swab | 0/19 | 0/9 |  |  |  |  | 0/28 | 0% |
| Serum | 0/19 | 0/9 |  |  |  |  | 0/28 | 0% |
| Urine | 4/17 | 0/9 |  |  |  |  | 4/26 | 15% |
| Total | 10/74 | 2/36 |  |  |  |  | 12/110 | 11% |
| All Farms Total | 34/419 | 2/198 | 0/223 | 1/245 | 0/152 | 0/72 | 37/1,309 | 2.8% |

<sup>1</sup>Number of days from BTM detection to individual cow sampling
<sup>2</sup>Quarter samples were taken from 20 individual sick cows and no individual quarters had IAV detected. Each individual was counted once as a non-detection;
<sup>1</sup>Number of days from farm bulk tank milk detection to sampling of cattle

**Table 4:**
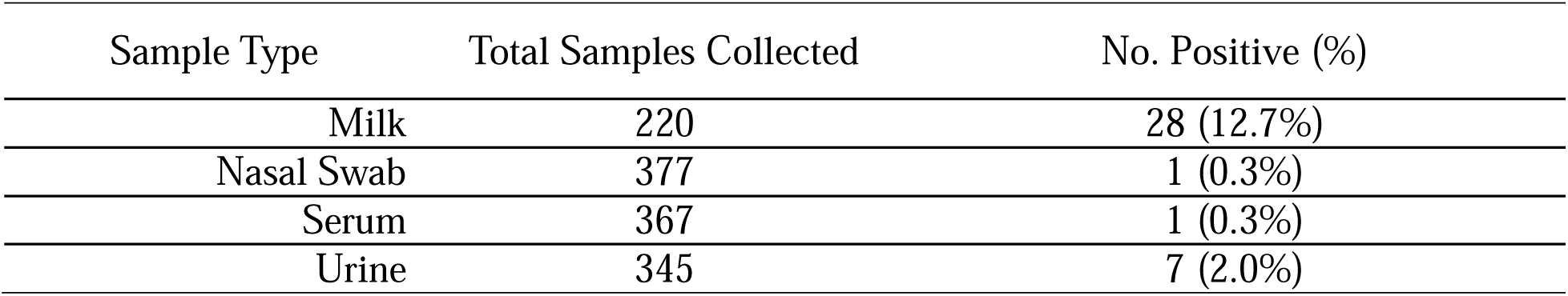
Summary of individual cow sample Influenza A virus PCR-positive test results for 6 California dairy herds, by sample type.

Serum samples collected from enrolled cattle were tested for IAV antibodies via IDEXX ELISA. The number of days from BTM detection to when these serum samples were collected can be seen in Table 3. Serum samples from two sick cows on Farm BM were ELISA positive. In addition, serum samples from two fresh cows on Farm BI and one sick cow on Farm BD were suspect ELISA positive. All other serum samples from tested cattle were ELISA negative. Viral RNA was found in milk samples from the cows with positive ELISA results on Farm BM. The single cow on Farm BD with suspect positive ELISA results had viral RNA found in multiple excretions (milk, urine and serum). The two fresh cows with suspect positive ELISA results on Farm BI did not have IAV detected in any excretion.

## DISCUSSION

This study details daily BTM and PLM IAV detections on dairy farms affected by the H5N1 virus. Because farms were enrolled prior to initial detection of IAV, trends in viral load (Ct values) measured from the beginning to the end of the outbreak period on each farm could be analyzed and visualized over time, giving estimates to the timeframe for dairy H5N1 outbreak management. Trends could be seen in the detection timelines on a majority of study farms; a Ct nadir was reached one to three weeks from first detection, and the time to the first test-negative day was typically between four and seven weeks.

Some farms (4/16; 25%) did not fit this trend, however, and had consistent daily detections of IAV for >75 days. BTM detections with a similarly prolonged time course have been seen in prior work looking at affected Colorado farms (USDA, 2025). Reasons for prolonged IAV BTM detection on any farm may be related to maintenance of the virus in certain farm environments (Stenkamp-Strahm et al., 2025), movement of naïve cows onto the operation, exposure of naïve cattle on the farm, or various approaches to the management/quarantine of sick cattle that may continue to shed infectious virus. A single herd with prolonged detection (Farm BF) also participated in the PLM study and had detections in just two pens of cattle at the end of the study period. This operation sends 5 to 6-month-old heifers offsite to be bred and raised until just prior to calving. One of the pens with prolonged detection was composed of cows that had recently calved, suggesting that potentially naïve cattle coming back to this operation may play a role in prolonged BTM detection.

The previous finding that IAV was present in BTM prior to the identification of sick cattle (NMPF, 2024) was confirmed in this study. Farms enrolled were made aware of their BTM detection status and likely anticipated that they would have clinically ill cattle in the days after detection. Regardless, clinical signs of H5N1 were seen anywhere from three to 14 days after initial detection on participating operations. *This information helps to validate BTM testing as an ideal method for early detection of this virus on dairy farms.* The work also supports the notion that sampling individual cattle is not a good early detection method of infected herds. A cross section of cattle from multiple cattle classes were tested for IAV early in the outbreak (within four to nine days of BTM detection) on six operations, and a very low percentage (< 3%) of H5N1 detections were made when looking across multiple collected sample types. On the day of individual cow sample collection, BTM Ct values for sampled farms ranged from 21 – 27.2 (data not shown). *In addition to supporting the notion that BTM testing is more sensitive than individual animal testing for virus early in an outbreak, this corroborates the likely presence of subclinical or nonclinical cows shedding viral RNA into their milk*.

For farms that submitted PLM samples, the H5N1 virus was detected in all pens within a relatively short time window (1-12 days) of clinical signs being detected in the herd. This timeframe is likely an overestimation; more than half of the farms in the PLM study started pen-level sampling when virus was already detected in all pens. When PLM was collected and tested near the peak of the outbreak (lowest achieved BTM Ct), Ct values from pens were often similar to those of the BTM measured on the same day, and measured nadirs were similar between samples from PLM and BTM. *Taken together, this information suggests that the virus, once on a farm, quickly spreads to all lactating pens of cattle.* A single farm in the study (Farm BC) submitted PLM samples prior to the date of herd clinical signs. On this farm, virus was detected in >30% of pens on the day clinical signs were first reported. *From a management perspective, this suggests that by the time clinical observations and segregation of sick cows is possible, the virus may have already spread considerably within a herd*.

Although our results suggest that intra-herd transmission of H5N1 is rapid, we still have much to learn regarding the transmission of this virus among dairy cattle. Results from intramammary infection experiments initially suggested contaminated milking equipment as a primary mode of transmission (Baker et al., 2024; Halwe et al., 2024), but infection using these methods has not been successfully reproduced in a lab setting (Lee et al., 2025). Although other dairy pathogens can be spread via contaminated milking equipment, history suggests that cow-to-cow transmission via this route in conventional milking systems may be relatively inefficient (Baxter et al., 1992; Deng et al., 2021), and unlikely to result in the proficient movement of virus observed. In addition to parlor surfaces and milking equipment, work by our group and others has shown H5N1 viral detection in dairy bioaerosols, wastewater, and the personal protective equipment of dairy personnel (Campbell et al., 2025; Stenkamp-Strahm et al., 2025). Individual cattle shed virus in excretions other than milk including whole blood, urine and nasal discharge (Lombard et al., 2025). Moreover, herds that had a relatively low incidence of disease during their H5N1 outbreak have shown high levels (>80%) of seroconversion (Pena Mosca et al., 2025; J. Lombard, Colorado State University, Fort Collins, CO, personal communication). Of the 33 early sampled cattle with IAV detection in this study, 7 (21%) had detection in urine, and 1 (3%) had detection in serum. *Findings from earlier studies, along with these results from individual cow and PLM sampling, leads to a specific hypothesis: some cows infected with H5N1 experience systemic infection or viremia, and there are likely multiple routes that cattle within a herd can be exposed to the virus*.

Results from a study that looked at the seroprevalence of influenza A in cattle across the US suggest that exposure to IAV may be more widespread in cattle than originally thought (Lang et al., 2025). Although all enrolled cattle were tested for the presence of serum IAV antibodies, only two tested cows on a single farm were ELISA positive, and these cows also had IAV detection in their milk samples. Compared to the other tested farms, this farm was sampled the greatest number of days (nine) after initial BTM detection. This fits with previous experimental studies that have shown the development of antibodies in serum and milk between seven and nine days after intramammary exposure (Baker et al., 2024; Halwe et al., 2024). Two fresh cows had serum antibody detection that was deemed to be ‘suspect’ when sampled seven days after BTM detection, and these individuals did not have IAV detected in any excretion. When heifer calves were experimentally aerosol inoculated with virus, they developed serum antibodies between seven and 13 days of exposure, and had sporadic IAV detections in nasal, oropharyngeal, ocular and saliva samples (Baker et al., 2024). For the operations enrolled in this study (and many others across the U.S) there had been no movement of infected cows prior to BTM detection, and we do not know how the H5N1 virus entered these herds. *It therefore remains unknown in a natural infection setting how many days after herd incursion that H5N1 may be detected in BTM samples, and how long after this incursion cattle may develop antibodies.* This timeframe would likely depend on a cow’s H5N1 exposure route, dose, and immune status.

One may consider the modification of BTM surveillance approaches, given the results from BTM and PLM detections in this study. For example, some herds may have outbreak windows that are less than or very close to one month in duration. *Monthly or more infrequent antigen-only testing may therefore miss herds that experience such short outbreak windows*. Also, when samples representing all milk produced by a herd on a single day were tested, some samples from a given herd would have IAV detected and others not. Herds in this work without these ‘simultaneous’ test result days were those that frequently submitted single daily samples (data not shown). *If using BTM for herd level detection surveillance or for quarantine release, there is a need to collect samples that represent all milking cows on an operation (i.e., samples representing all milk shipped on a given day) for best detection sensitivity.* Further, in this study there were days when pen-level samples from a herd had IAV detection, while BTM samples from the same day were determined to be test-negative. This was observed toward the end of farm outbreak periods when results were trending toward non-detection, *supporting a notion that the sensitivity of BTM IAV testing decreases as viral load decreases*. At the time of this writing, very little has been published regarding the detection sensitivity of IAV in BTM or PLM samples. Future work should focus on measuring IAV detection sensitivity in milk, especially in cases where only a few animals may be contributing low levels of viral RNA to an aggregated sample.

### Limitations

Farms enrolled in the current work were instructed to submit a single milk sample from each bulk tank that was shipped daily. This was in an attempt to represent all lactating cows on an operation, since cows may be milked multiple times and milk from multiple bulk tanks may be shipped each day, depending on farm size. The number of samples submitted compared to the number of bulk tanks shipped was not monitored, however, so the authors are limited in knowing if all cows on each operation were truly represented in Ct measures, or whether duplicate samples from shipped tanks were submitted. We also cannot extrapolate viral loads in the milk from a given tank or pen of animals to how many individual cows may be shedding virus. This limits our understanding as to the exact speed of intra-herd viral spread; although virus was detected in pens relatively quickly, it’s possible that only a few individuals were contributing viral RNA to collected samples. Comparing the magnitude of each individual farms’ outbreak is also not possible given BTM Ct values alone. Standard curves are not routinely completed by NAHLN labs on samples submitted for outbreak testing, but a Ct nadir value of 18 (the lower end of the range seen in this work) would likely equate to ∼1,900,000 viral copies of RNA, while a Ct value of 21 (the upper end of the range seen in this work) would equate to ∼228,000 copies (S. Robbe-Austerman, NVSL, Ames, IA, personal communication).

We currently lack a case definition for H5N1 disease in dairy cattle. What may deem a cow ‘clinical’ for H5N1 is therefore variable across farms that have been affected by this virus to date, including farms enrolled in the current work. For the individual cow study, the way sick cattle were presented to LVC personnel was not uniform across operations and may represent a study bias in the percentages of individual cow IAV detected by operation. As an example, two of the six farms (Farm BN and Farm BD) used biometric data systems, so may have had an increased sensitivity for detecting cows potentially infected with H5N1 compared to the other operations studied. All farms were sampled from 2 days prior to 6 days after personnel-deemed detection of clinical signs, with most sampling occurring within 1-3 days of clinical signs and a low number of purported H5N1 cases having been identified on each farm when sampling took place.

### Conclusion

This work has described daily detections of IAV in BTM and PLM samples from multiple dairies affected by H5N1. Our findings describe both trends and variabilities in aggregate milk detections over time, showing the highest aggregate milk viral loads occurring within three weeks of initial detection and overall detections of IAV waning by two months. This work also confirms a need to study farm-level factors that play a role in prolonged detection of the virus, as a subset of herds did not fit these trends. Although this work has not clarified transmission routes of the virus between cows, PLM detections suggest that intra-herd spread of the virus is relatively quick, especially considering the clinical disease onset in each herd and the overall low number of detections made from individual cow excretions. The authors recommend completing analyses that focus on IAV rRT-PCR detection sensitivities in aggregate milk, given the detection of virus in PLM samples when BTM from the same farms had become test-negative.

Our results support the current use of BTM testing for early detection of dairy H5N1 outbreaks. The patterns of herd viral detection seen here should be used to guide future surveillance approaches. Specific recommendations to improve the effectiveness of current monitoring include testing BTM at intervals of less than one month, so as not to ‘miss’ identifying infected farms. Additionally, testing aggregate milk samples that represent all cows currently lactating on an operation should be pursued, given that study farms had BTM samples with detection and non-detection on a single day.

## Data Availability

All data produced in the present work are contained in the manuscript or available online at the following DOI: 10.17605/OSF.IO/TFJ87

https://doi.org/10.17605/OSF.IO/TFJ87

## Abbreviations

CS: clinical signs of H5N1 disease in cows,
H5N1: highly pathogenic avian influenza virus of the H5N1 subtype,
IAV: influenza A virus,
BTM: bulk tank milk,
PLM: pen level milk

## SUPPLEMENTAL MATERIAL

The data supplement can be found at the following DOI: 10.17605/OSF.IO/TFJ87

## Acknowledgements

The authors would like to thank the following personnel of Lander Veterinary Clinic for their work collecting and characterizing samples: Yvette Guzman, Ernesto Padilla, Alondra Ramirez, Carina Montañez, Jessica Pacheco, and Giselle Leon. The authors would like to additionally thank the Iowa State University Veterinary Diagnostic Laboratory team for their testing of aggregate milk and individual cow samples. We lastly would like to thank the California Department of Food and Agriculture for their collaboration and cooperation during this work. *Ethics statement* The Colorado State University Institutional Animal Care and Use Committee (IACUC) determined this study to be exempt from IACUC approval. CSU IACUC #5980.

## Artificial intelligence disclosure

Artificial intelligence (AI) was not used in the writing of any section of the manuscript body. During the preparation of this work, the authors used ChatGPT to generate the interpretive summary, reviewed and edited the content, and take full responsibility for that section of the manuscript.

## REFERENCES

Baker, A. L., B. Arruda, M. V. Palmer, P. Boggiatto, K. Sarlo Davila, A. Buckley, G. Ciacci Zanella, C. A. Snyder, T. K. Anderson, C. Hutter, T. Q. Nguyen, A. Markin, K. Lantz, E. A. Posey, M. K. Tor-chetti, S. Robbe-Austerman, D. R. Magstadt, and P. J. Gorden. 2024. Experimental reproduction of viral replication and disease in dairy calves and lactating cows inoculated with highly pathogenic avian influenza H5N1 clade 2.3.4.4b. Nature. 10.1101/2024.07.12.603337.

Baxter, J. D., G. W. Rogers, S. B. Spencer, and R. J. Eberhart. 1992. The effect of milking machine liner slip on new intramammary infections. J. Dairy Sci. 75:1015–1018. 10.3168/jds.S0022-0302(92)77844-8.

CDFA (California Department of Food and Agriculture). 2024. Healthy Dairy Cattle H5N1 Bird Flu Testing Protocols for Monitored Herd, Pre-Movement, and Non-Monitored Herd Surveillance for Producers. Accessed Jul. 2025. https://www.cdfa.ca.gov/AHFSS/Animal_Health/docs/surveillance_testing_of_dairy_cattle_protocol-for_producers.pdf.

Campbell AJ, M. Shephard, A.P. Paulos, M. Pauly, M. Vu, C. Stenkamp-Strahm, K. Bushfield, B. Hunter-Binns, O. Sablon, E.E. Bendall, W.J. Fitzimmons, K. Brizuela, G. Quirk, N. Kumar, B. McCluskey, N. Shetty, L.C. Marr, J.J. Guthmiller, K. Abernathy, A.S. Lauring, B.T. Melody, M. Wolfe, J. Lombard, S.S. Lakdawala. 2025. Surveillance on California dairy farms reveals multiple sources of H5N1 transmission. bioRxiv. 2025.07.31.666798. 10.1101/2025.07.31.666798

Caserta, L. C., E. A. Frye, S. L. Butt, M. A. Laverack, M. Nooruzzaman, L. M. Covalenda, A. Thompson, M. Prarat Koscielny, B. Cronk, A. Johnson, K. Kleinhenz, E. E. Edwards, G. Gomez, G. R. Hitchener, M. Martins, D. R. Kapczynski, D. L. Suarez, E. R. Alexander Morris, T. Hensley, J. S. Beeby, M. Lejeune, A. Swinford, F. Elvinger, K. M. Dimitrov, and D. G. Diel. From birds to mammals: Spillover of highly pathogenic avian influenza H5N1 virus to dairy cattle led to efficient intra- and interspecies transmission. Nature. 10.1101/2024.05.22.595317.

CDA (Colorado Department of Agriculture). 2024. Order of the commissioner of agriculture – statewide mandatory bulk tank HPAI testing. Accessed Dec. 20, 2024. https://drive.google.com/file/d/17KS5LLEG-Dir1s9fpGVL8Mnp62ACCz2a/view.

Deng Z, G. Koop, H. Hogeveen, E.A.J. Fischer, B.H.P Van den Borne, R. van der Tol, T.J.G.M. Lam. 2021. Transmission dynamics of Staphylococcus aureus and Streptococcus agalactiae in a Dutch dairy herd using an automatic milking system. Prev. Vet. Med. 192:105384. 10.1016/j.prevetmed.2021.105384

Gilbertson, B., and K. Subbarao. 2023. Mammalian infections with highly pathogenic avian influenza viruses renew concerns of pandemic potential. J. Exp. Med. 220:e20230447. 10.1084/jem.20230447.

Graziosi, G., C. Lupini, E. Catelli, and S. Carnaccini. 2024. Highly pathogenic avian influenza (HPAI) H5 clade 2.3.4.4b virus infection in birds and mammals. Animals (Basel) 14:1372. 10.3390/ani14091372.

Halwe, N. J., K. Cool, A. Breithaupt, J. Scho n, J. D. Trujillo, M. Nooruzzaman, T. Kwon, A. K. Ahrens, T. Britzke, C. D. McDowell, R. Piesche, G. Singh, V. Pinho Dos Reis, S. Kafle, A. Pohlmann, N. N. Gaudreault, B. Corleis, F. M. Ferreyra, M. Carossino, U. B. R. Balasuriya, L. Hensley, I. Morozov, L. M. Covaleda, D. Diel, L. Ulrich, D. Hoffmann, M. Beer, and J. A. Richt. 2025. H5N1 clade 2.3.4.4b dynamics in experimentally infected calves and cows. Nature 637:903–912. 10.1038/s41586-024-08063-y.

Lang Y., L. Shi, D. Gupta, C. Dai, M. A. Khalid, M.Z. Zhang, X. Wan, R. Webby, W. Ma. 2025. Detection of antibodies against influenza A viruses in cattle. J. Virol. e0213824. 10.1128/jvi.02138-24

Lee C., N.N. Tarbuck, H.J. Cochran, B.M. Foreman, P. boley, S. Khatiwada, A. Dhakal, K.O. Adefaye, J. Schrock, M.J. Jahid, T. Laocharoensuk, R. Suresh, O. Shekoni, E. Stevens, S. Dolatyabi, C. Sanders, E. Ohl, D. Huey, J. Hanson, R. Gourapura, R.J. Webby, C.J. Warren, S.P. Kenney, A.S. Bowman. 2025. Dairy cows infected with influenza A(H5N1) reveals low infectious dose and transmission barriers. Res. Gate. 10.21203/rs.3.rs-6900680/v1

Lombard J., C. Stenkamp-Strahm, B. McCluskey, B. Melody. 2025. Evidence of viremia in dairy cows naturally infected with influenza A virus, California, USA. Emerg. Infect. Dis. 31:1425– 1427. 10.3201/eid3107.250134

NMPF (National Milk Producers Federation). 2024. HPAI H5N1 Virus Spillover into Dairy Cattle. Accessed Jun. 10, 2024. https://www.nmpf.org/wp-content/uploads/2024/11/NMPF_H5N1-virus-early-detection-bulk-tank-milk.pdf.

Nobrega D.B., J.E. French, D.F. Kelton. 2023a. A scoping review of the testing of bulk milk to detect infectious diseases of dairy cattle: diseases caused by bacteria. J. Dairy Sci. 106:1986– 2006. 10.3168/jds.2022-22395.

Nobrega D.B., J.E. French, D.F. Kelton. 2023a. A scoping review of the testing of bulk tank milk to detect nonbacterial pathogens or herd exposure to nonbacterial pathogens in dairy cattle. J. Dairy Sci. 106:5636–5658. 10.3168/jds.2022-22586.

Peña-Mosca, F., E.A. Frye, M.J. MacLachlan, A.R. Rebelo, P.S.B. de Oliveira, M. Nooruzzaman, M.P. Koscielny, M. Zurakowski, Z.R. Lieberman, W.M. Leone, F. Elvinger, D.V. Nydam, D.G. Diel. 2025. The impact of highly pathogenic avian influenza H5N1 virus infection on dairy cows. Nat. Commun. 16:6520. 10.1038/s41467-025-46520-4.

Spackman, E., D. R. Jones, A. M. McCoig, T. J. Colonius, I. V. Goraichuk, and D. L. Suarez. 2024a. Characterization of highly pathogenic avian influenza virus in retail dairy products in the US. J. Virol. 98:e00881–24. 10.1128/jvi.00881-24.

Stenkamp-Strahm, C., B. MucCluskey, B. Melody, B. Christiansen, N. Urie, N. Amey, R. Lomkin, A.J. Campbell, S.S Lakdawala, J. Lombard. 2025. Dairy environments with milk exposure are most likely to have detection of influenza A virus. medRxiv. 2025.09.03.25335023. 10.1101/2025.09.03.25335023.

Tammiranta, N., M. Isomursu, A. Fusaro, M. Nylund, T. Nokireki, E. Giussani, B. Zecchin, C. Terregino, and T. Gadd. 2023. Highly pathogenic avian influenza A (H5N1) virus infections in wild carnivores connected to mass mortalities of pheasants in Finland. Infect. Genet. Evol. 111:105423. 10.1016/j.meegid.2023.105423.

USDA. 2024a. HPAI confirmed cases in livestock. USDA-Animal and Plant Health Inspection Service (APHIS). Accessed Dec. 6, 2024. https://www.aphis.usda.gov/livestock-poultry-disease/avian/avian-influenza/hpai-detections/hpai-confirmed-cases-livestock.

USDA. 2024b. Federal order requiring testing for and reporting of highly pathogenic avian influenza (HPAI) in livestock. Accessed Jun. 10, 2024. https://www.aphis.usda.gov/sites/default/files/dairy-federal-order.pdf.

USDA. 2024c. USDA announces new federal order, begins national milk testing strategy to address H5N1 in dairy herds. Accessed Dec. 20, 2024. https://www.usda.gov/article/usda-announces-new-federal-order-begins-national-milk-testing-strategy-address-h5n1-dairy-herds.

USDA. 2025. Technical Brief: Estimating the Probability of Highly Pathogenic Avian Influenza Detection in Bulk Tank Milk at Various Sampling Times Using Disease Event Surveillance Data. Accessed Aug. 8, 2025. https://www.aphis.usda.gov/sites/default/files/tech-brief-hpai-btm-surveillance.pdf

Utah Department of Agriculture and Food (UDAF). 2024. UDAF enacts mandatory surveillance of HPAI in Cache County dairies. Oct. 24, 2024. Accessed Aug. 15, 2025. https://ag.utah.gov/2024/10/23/udaf-enacts-mandatory-surveillance-of-hpai-in-cache-county-dairies/.

